# Factors Influencing the Effectiveness of AI-Assisted Decision-Making in Medicine: A Scoping Review

**DOI:** 10.1101/2025.09.02.25334863

**Authors:** Nicholas J Jackson, Katherine E. Brown, Rachael Miller, Matthew Murrow, Michael R Cauley, Benjamin X Collins, Laurie L Novak, Natalie C Benda, Jessica S Ancker

## Abstract

**Objective:** Research on artificial intelligence-based clinical decision-support (AI-CDS) systems has returned mixed results. Sometimes providing AI-CDS to a clinician will improve decision-making performance, sometimes it will not, and it is not always clear why. This scoping review seeks to clarify existing evidence by identifying clinician-level and technology design factors that impact the effectiveness of AI-assisted decision-making in medicine.

**Materials and Methods:** We searched MEDLINE, Web of Science, and Embase for peer-reviewed papers that studied factors impacting the effectiveness of AI-CDS. We identified the factors studied and their impact on three outcomes: clinicians’ attitudes toward AI, their decisions (e.g., acceptance rate of AI recommendations), and their performance when utilizing AI-CDS.

**Results:** We retrieved 5,850 articles and included 45. Four clinician-level and technology design factors were commonly studied. Expert clinicians may benefit less from AI-CDS than non-experts, with some mixed results. Explainable AI increased clinicians’ trust, but could also increase trust in incorrect AI recommendations, potentially harming human-AI collaborative performance. Clinicians’ baseline attitudes toward AI predict their acceptance rates of AI recommendations. Of the three outcomes of interest, human-AI collaborative performance was most commonly assessed.

**Discussion and Conclusion:** Few factors have been studied for their impact on the effectiveness of AI-CDS. Due to conflicting outcomes between studies, we recommend future work should leverage the concept of ‘appropriate trust’ to facilitate more robust research on AI-CDS, aiming not to increase overall trust in or acceptance of AI but to ensure that clinicians accept AI recommendations only when trust in AI is warranted.

## INTRODUCTION

Given the increasing effectiveness of artificial intelligence (AI) for diagnostic and clinical reasoning tasks^1–5^, many studies have evaluated the potential for these systems to augment clinical decision-making via the use of AI-based clinical decision support (AI-CDS), with promising results^6–9^. However, recent studies have found that AI-CDS may not improve clinicians’ decision-making performance, even when the AI-CDS performs well (e.g., has high diagnostic accuracy). For example, Goh et al.^10^ found that providing physicians with access to a large language model (LLM) chatbot assistant did not improve physicians’ performance on a diagnostic reasoning task, even though the chatbot performed better on this task than physicians did. Similarly, Yu et al.^11^ found that providing AI-CDS for chest X-ray diagnosis did not improve on clinicians’ diagnostic performance. Similar results have been observed in real-world AI-CDS deployment: when AI-CDS recommendations conflict with clinicians’ initial judgment, clinicians reject these recommendations, leading to no change in patient outcomes^12,13^. These studies demonstrate the lack of clarity in this field: sometimes providing AI-CDS to a clinician will improve their decision-making performance^6,7,9^, sometimes it will not^10–12^, and it is not yet clear why. The benefits of AI-CDS for patient outcomes cannot be realized without a more comprehensive understanding of how the design features of the AI-CDS, characteristics of the clinician, and aspects of the clinical context influence the success or failure of AI-assisted decision-making.

Although insights are available from the existing research on non-AI (i.e., traditional) CDS systems,^14–17^ they are not sufficient to resolve the questions about AI-CDS. This is largely because the complexity and opacity of AI-CDS are significantly higher than those of traditional CDS and, as a result, users may be less willing to trust or accept the recommendations of AI-CDS^14,18^.

Additionally, an emerging body of literature attempts to improve AI-assisted decision-making by modifying how users interact with AI systems through new computational and design approaches^23–25^. Promising approaches include providing explanations for AI decisions^26^ (explainable AI) or disclosing AI uncertainty to the decision-maker^27,28^ (uncertainty quantification). Despite the promise of these approaches, relatively little of this research has studied medical decision-making, which carries unique ethical, legal, and social implications^15,16,23–25,29^. As a result, it is not yet clear how these findings apply to medical AI.

Given the rapid growth of medical AI and the mixed results of AI-CDS in both experimental and real-world settings, we conducted a scoping literature review to identify technology design and clinician-level factors that influence the effectiveness of human-AI collaborative decision-making in medicine. We focused on three important outcomes relevant to effectiveness of decision support: clinicians’ attitudes toward AI, rate of acceptance of AI-CDS recommendations, and human-AI collaborative performance on a task of interest (e.g., differential diagnosis).

## METHODS

### Review Methodology

We conducted our review in accordance with the PRISMA-ScR guidelines^30^ (Supplementary Materials, Table 1). Informed by consultation with an experienced medical librarian, we searched MEDLINE, Web of Science, and Embase for peer-reviewed papers published in English between June 1^st^ 2013 (the end date of a related review)^14^ and May 1^st^ 2025 (the date the search was conducted). Because this research area does not use consistent terminology^15,16,23,24^, we developed our search query by manually identifying 20 relevant articles that met our inclusion criteria and then adjusted our query until it identified all 20. This query ensured that the title or abstracts of studies included: a term about AI (e.g., artificial intelligence, machine learning, etc.), a term about medicine (e.g., diagnose, patient, medicine, etc.), and a term about assisted decision-making (e.g., decision-support, recommendation, human-AI, etc.). Full search queries are available in the Supplementary Materials and an Open Science Framework repository (https://osf.io/un32b/).

We leveraged the literature review tool Covidence^31^ to screen articles, perform full-text review, and extract data. Titles and abstracts were screened by one reviewer, who maximized recall with loosened inclusion criteria (i.e., including any study where the title or abstract mentioned any human interaction with AI in a medical context). Full-text articles were then screened by 2 reviewers according to these inclusion criteria:

1. Studies must have conducted an experiment or observed an actual implementation of AI-CDS and collected data on participant use of AI-CDS. We defined an AI-CDS system as a CDS system with a decision-making process that relies on data-driven learning (e.g., machine learning) or pattern recognition, beyond explicitly programmed logic. This criterion enabled us to include sophisticated decision-making systems of varying types, while capturing the key attributes of AI-CDS (i.e., opacity and complexity) that differentiate them from existing CDS systems that have been well-studied elsewhere.
2. Study participants must be healthcare professionals with clinical knowledge relevant to the decision-making task under study.
3. Studies must have assessed either: an objective measure of a user’s actions (e.g., the decision to accept or reject a recommendation from AI-CDS); a subjective measure of a user’s attitudes towards the AI-CDS (e.g., their trust in the AI-CDS); or their performance with AI-CDS (e.g., accuracy or area under the receiver operating characteristics curve).
4. Studies must have measured or manipulated an additional variable to determine its effect on either the clinicians’ attitudes towards the AI-CDS, their decisions/actions when using the AI-CDS, or their performance when using the AI-CDS. These three measures were selected a-priori as they align with common concepts from several theoretical frameworks for technology-assisted decision-making^32–34^.
5. Studies must have included at least 30 decision-makers. This cutoff was chosen for two reasons. First, we were primarily interested in participant-level effects (e.g., the impact of explainable AI on clinician trust). While studies with fewer than 30 participants could be well-powered at the level of individual decisions, they are unlikely to return generalizable results at the participant-level. Second, it represents a pragmatic cutoff to limit the number of articles needed to review.

All disagreements between reviewers were settled via consensus.

We extracted information on study design and participants, which decision-making or performance outcomes were studied (i.e., attitudes, actions, and performance), how these outcome variables were operationalized, and what factors were studied for their impact on these outcomes. When measuring performance, we used the primary performance measure reported by the authors of each study as each application of AI-CDS has its own unique objectives and performance criteria. Additionally, to enable more granular analysis, we differentiated the performance of clinicians alone from clinicians using AI-CDS (hereafter, human-AI collaborative performance) and from the AI-CDS alone. Data extraction was initially performed by a single author. After the initial data extraction was completed, this process was repeated (by the same author). Differences between extraction rounds were then reconciled on an item-by-item basis within Covidence. Extracted information was processed via a custom Python script (manually created by the study team) to clean and map free-text inputs onto the discrete categories we used for our analysis. The protocol for this study was registered with the Open Science Framework. The raw extracted information, code, processed data, and citations are provided in an Open Science Framework repository (https://osf.io/un32b/).

## RESULTS

### Screening

We identified 10,027 articles, 4,177 of which were removed as duplicates. The title and abstract screening excluded 5,090 articles, and the full-text screening phase removed 715 articles, leaving 45 articles for inclusion in our study (Figure 1). The most common exclusion reasons at the full-text stage were due to insufficient numbers of participants (N=222 studies) and the lack of an independent variable that was assessed for its impact on human-AI interaction (N=209). The full list of included studies is included in Supplementary Materials, Table 2.

**Figure 1:**
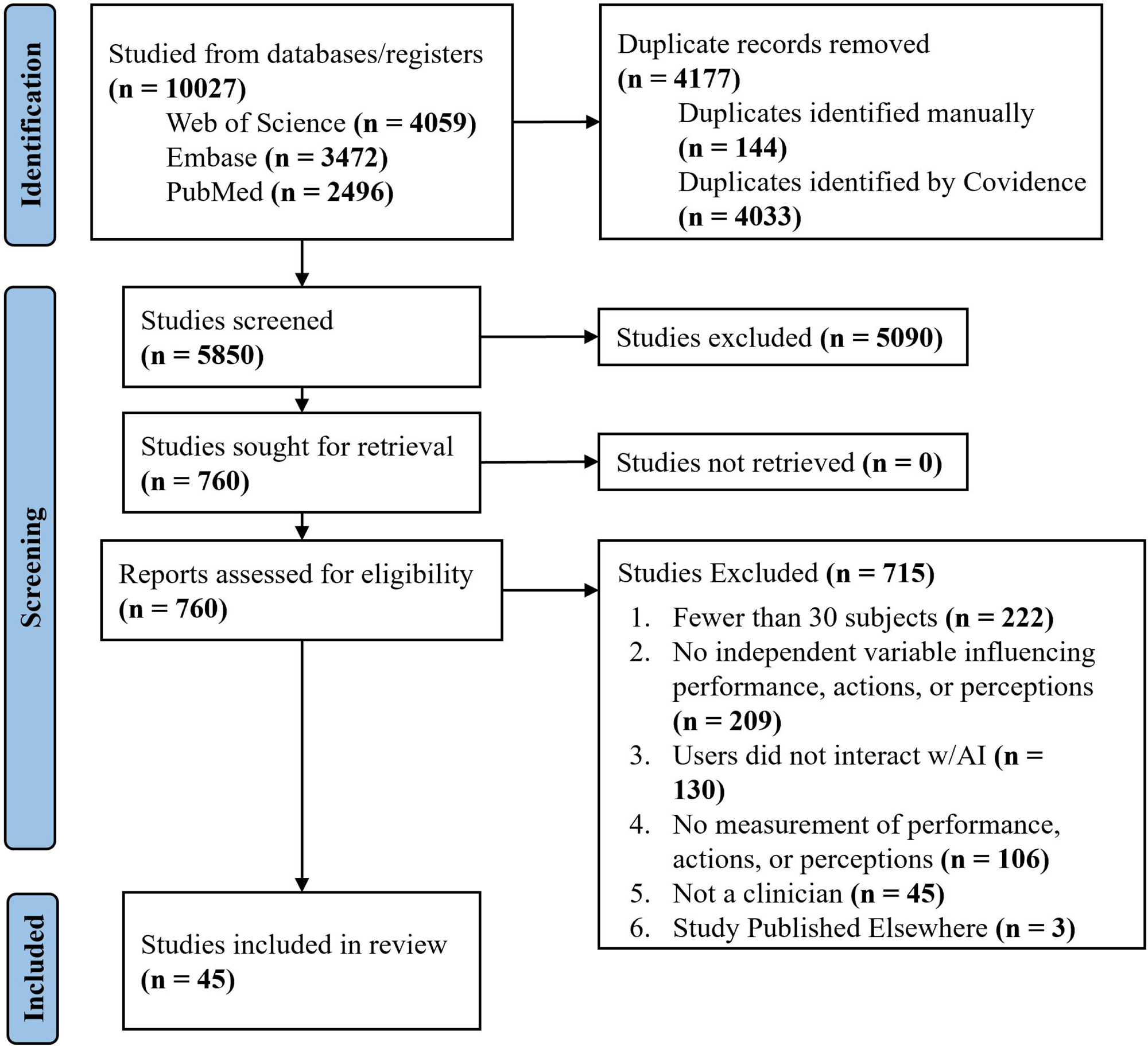
PRISMA diagram for study inclusion.

### Study Designs & Sample Sizes

The number of decision-making participants across studies was skewed with a mean of 161.4 participants and a median of 86.0, largely due to a small number of studies that had over 250 participants^7,37–43^ (Figure 2). Most studies used image or video-based modalities (N=29) with the rest using clinical vignettes (N=12) or electrocardiograms (N=4). Studies primarily recruited attendings (N=42) or physician trainees such as interns, residents, or fellows (N=24). A smaller number of studies recruited nurses (N=3) and advanced practice providers (N=3) with registered nurse anesthetists, mental health professionals, ambulance staff, biochemistry staff, and medical students each being recruited in one study. Studies were primarily conducted via surveys (N=26) or in simulated laboratory settings (N=17) with few studies conducted in real-world clinical environments (N=2). The AI systems used were primarily deep learning (N=26), but interestingly 12 studies used simulated AI systems, where the advice provided to users was controlled by the study designers. Of the remaining studies several used large language models (N=3) and traditional machine learning (N=2) and in two studies it was unclear what AI system was used.

**Figure 2:**
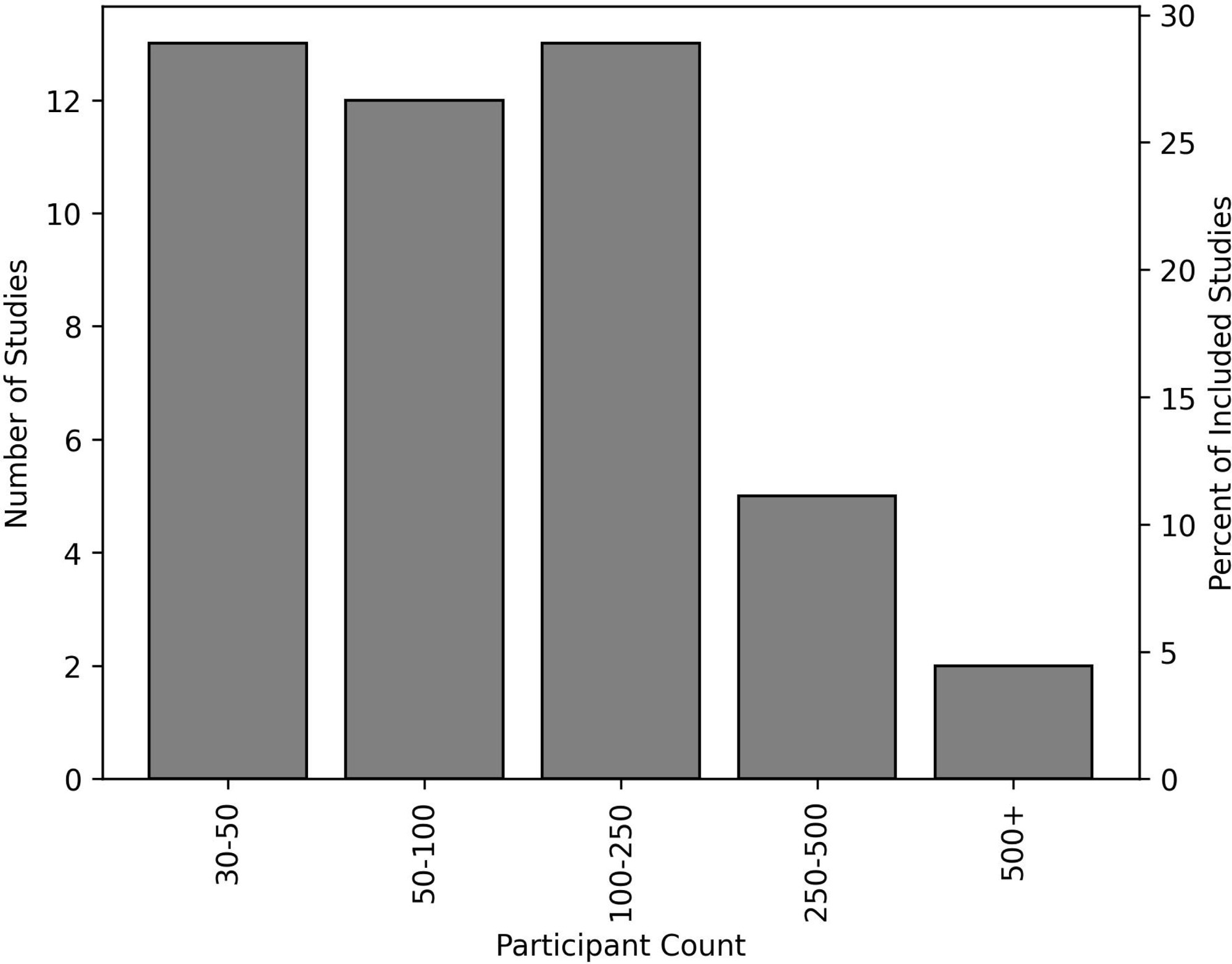
Participant count in the included studies.

Most studied investigated diagnostic tasks (N=36) with a few others making decisions about drug prescription (N=3) and triage/risk assessment (N=3). The remaining three studies each investigated unique tasks which we categorized as: screening, surgical guidance, and determining whether to call emergency dispatch.

### Outcomes Assessed in the Included Studies

Participants’ performance with AI-CDS was the most studied outcome (N=33, 73% of studies), followed by participant attitudes (N=20, 44%) and actions (N=18, 40%). Importantly, few studies measured all three of these outcomes (N=5, 11%). Participants’ performance was primarily measured via accuracy (e.g., percentage of decisions that were correct, N=26/33), followed by area under the receiver operating characteristic curve (N=4/33), sensitivity (N=1/33), rate of adherence to clinical guidelines (N=1/33), and a custom score provided by subject-matter experts (N=1/33).

In the 20 studies assessing clinician attitudes towards AI, trust was the most commonly studied clinician attitude (N=11/20). This was followed by a series of indirect measures of clinicians’ attitudes toward AI-CDS. Namely, clinicians’ confidence either in the AI-CDS or their AI-assisted decisions (N=8/20), clinicians’ perception of the AI-CDS’ utility or quality (N=7/20), and clinicians’ self-reported understanding of the AI-CDS (N=4/20).

Participants’ actions were most commonly measured via the agreement between the clinician and the AI-CDS (N=8/18). The next most common measure of users’ actions was decision-switching (N=7/18) which was broken down into: (1) the rate at which participants changed a binary decision (e.g., a diagnosis) after receiving advice from AI-CDS (N=5/18) and (2) weight-on-advice^44^, a weighted measure of how much a participant changed their decision about a continuous quantity (e.g., selecting a medication dose), (N=2/18). Additionally, two studies measured the participants’ level of agreement with AI-CDS when the AI-CDS provided incorrect advice, which was referred to as ‘over-reliance’ or ‘automation bias’.

### Factors Impacting the Effectiveness of AI-Assisted Decision-Making

A summary of which factors were assessed by which studies for their impact on AI-Assisted decision-making is summarized in Figure 3.

**Figure 3:**
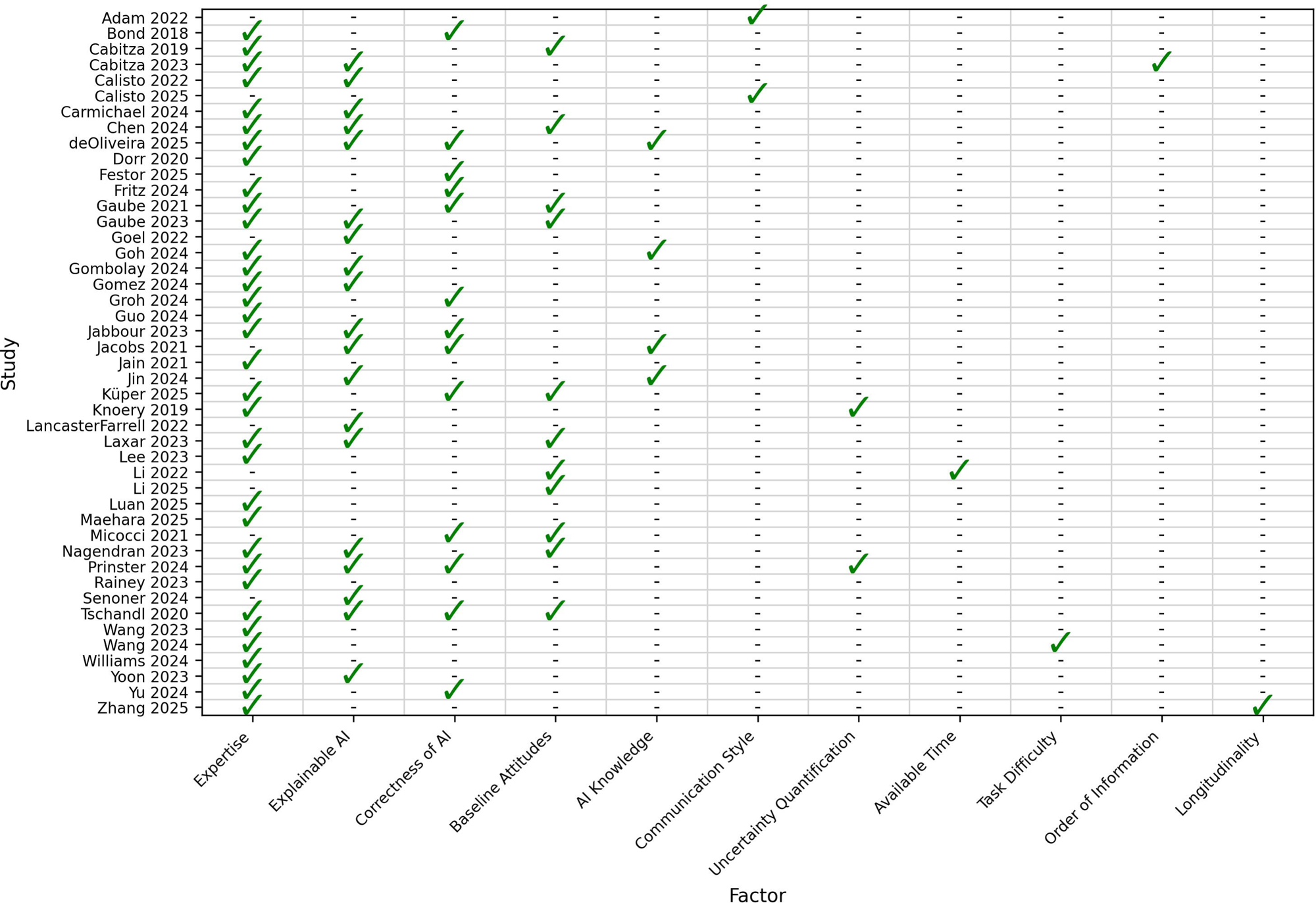
Summary of variables studied in each AI-assisted decision-making study.

#### Expertise

The most frequently studied variable was expertise (i.e., whether the clinician using the AI system was an expert in the clinical domain under study) (N=34). Which individuals were considered experts differed in each study as these studies covered many different clinical domains. Typically, experts were distinguished from non-experts via their years of experience or their level of clinical training (e.g., subspecialist vs attending vs resident vs medical student).

Nine studies found that AI assistance improved performance for experts and non-experts to a similar degree,^7,10,11,38,45–49^ two found that AI improved performance more for experts,^43,50^ and six found that AI improved performance more for non-experts^42,51–55^.

Of the five studies measuring the impact of expertise on clinicians’ attitudes toward AI, four found that experts perceived AI as less helpful or exhibited lower trust in it than non-experts^56–59^. However, an additional study suggested no relationship between expertise and trust^60^.

In the 11 studies that assessed the impact of expertise on clinicians’ actions, the effect was mixed. Four studies found that experts agreed with or aligned their decisions to AI-CDS recommendations less often than non-experts^42,47,51,56^, but seven studies found no clear relationship between participant expertise and their actions with AI^61–63^.

Several studies additionally evaluated how expertise interacted with other variables such as explainable AI (xAI) or incorrect advice. Specifically, in one study experts rated xAI as lower quality than non-experts^64^ Another study found that those experts who did find xAI useful (i.e., explainable^65^) exhibited decreased performance^40^ while two studies reported that experts were less likely to follow incorrect advice than non-experts^39,51^.

#### Explainable AI

The next most studied factor was the use of xAI to better communicate the AI-CDS’ decision-making process to clinicians (N=19). We defined xAI broadly as any AI system containing a component that “aims to increase the transparency, trustworthiness and accountability of the AI system”^66,67^. Therefore, any approach that the authors described as intending to explain, support, or clarify AI-CDS decisions was considered xAI for the purposes of this study. Notably, there were mixed effects of xAI on performance. Of the 13 such studies, five studies found that xAI improved performance^55,58,64,68,69^, but six identified no effect^40,41,52,70–72^, and two showed that xAI worsened human-AI collaborative performance compared to human-AI performance with non-explainable AI^42,57^.

This heterogeneous effect of xAI on performance was mirrored by the effect of xAI on clinicians’ attitudes toward AI-CDS (N=10). In terms of trust-related attitudes, four studies identified that xAI could increase^57,62,64,70^ such attitudes with three showing a decrease^58,60,73^, and three others showing no effect^40,69,72^. However, these results were often observed under different circumstances. For example, xAI decreased clinicians’ trust and understanding of the AI when it ‘explained’ incorrect advice^60,73^, which is a desirable outcome as it potentially decreases acceptance of incorrect AI advice. Similarly, two studies found that xAI increased clinicians’ acceptance of correct AI advice, thereby increasing human-AI collaborative performance^64,68^.

However, these beneficial effects of xAI were not observed in all studies. In three studies, xAI was more likely to decrease participants’ performance when the AI-CDS was wrong (i.e., it convinced users to accept incorrect AI recommendations more frequently than with non-explainable AI)^68,69,72^.

Additionally, four studies compared the type of xAI used. When comparing local explanations vs global explanations (i.e., explanations about the model’s architecture and overall performance), local explanations (i.e., decision-specific or patient-specific) were more effective, increasing performance^69^ and agreement with AI advice^61^. However, two additional studies compared different local xAI approaches, finding no differences in performance^40,58^.

#### AI Correctness

Thirteen studies stratified their analyses on whether the advice from the AI-CDS was correct or incorrect. Unsurprisingly, these studies often found that incorrect AI-CDS advice decreased diagnostic performance^11,38,39,41,42,69,72,74,75^. However, participants showed some resilience to this as incorrect advice decreased users’ trust^60^ in the AI-CDS and decreased their confidence in their decisions^69,75^ (with one exception^72^). Moreover, participants rated correct advice as being more useful^39,69^ than incorrect advice and agreed with incorrect AI recommendations less frequently than correct ones^47,76^. In particular, both experts^39,51^ and those with higher self-confidence^74^ were less susceptible to following incorrect AI advice.

#### Baseline Attitudes Towards AI

Eleven studies assessed users’ baseline attitudes about the AI-CDS or AI in general before they interacted with the AI-CDS. The attitudes measured differed widely with most studies reporting some measure of trust in AI^47,62,63,77^ or users’ confidence in their own ability (sometimes relative to AI)^42,47,61,74,78^, with several studies inferring attitudes toward AI by comparing acceptance of AI advice to human advice^39,56,64^. No studies assessed the impact of baseline attitudes on performance. Participants with negative baseline attitudes towards AI were less likely to agree with AI recommendations in two studies^47,77^. Similarly, in two separate studies such negative attitudes caused participants to accept advice more frequently when told that it came from another human^56,64^. Conversely, one study found that those with higher trust in technology agreed with AI-CDS more frequently^47^. Lastly, only one study found that baseline attitudes toward AI did not impact decision-making^63^.

#### Emerging Evidence

A few factors were assessed by only one or two studies each. For example, two studies assessed the impact of conveying the AI-CDS’ level of certainty to participants (e.g., the probability of the AI-CDS’ decision being correct). One such study found that this increased participants’ diagnostic performance; however, clinicians’ performance decreased when the AI-CDS conveyed that it was highly certain, possibly indicating automation bias^79^. The other such study observed different decision-making patterns between experts and non-experts when AI was uncertain.

Specifically, with low-certainty AI recommendations, experts benefitted from xAI but non-experts did not^69^. Moreover, several studies assessed participants’ knowledge of AI, finding that educational interventions did not impact participants’ trust in AI^60^, but sharing performance metrics of the AI-CDS increased clinicians’ trust^70^. Another study found that users who were more familiar with AI rated AI-CDS as having higher utility^72^. However, these studies found no effect^10^ or a detrimental effect^72^ of AI knowledge on decision-making performance.

Two studies assessed AI-CDS communication styles. Specifically, one study found that providing prescriptive advice (i.e., instructions on what to do) increased agreement with AI compared to descriptive advice (i.e., neutrally conveying information)^37^. Another study found that a similar strategy to prescriptive advice (which the authors refer to as assertive advice) had a positive effect on some measures of clinicians’ attitudes towards the AI-CDS^80^. One study assessed the order in which AI recommendations were provided to participants (i.e., before vs after making their own decision about the patient). They observed that human-AI collaborative performance was higher when AI recommendations were presented before the clinician made their own assessment of the patient^52^.

Participants’ actions with AI were found to evolve over time; one study observed that clinicians’ adherence to clinical guidelines increased continuously over a two-week AI-CDS intervention period^81^. Lastly, applying time restrictions to participants was found to hinder their diagnostic performance without AI, but when AI-CDS was available, participants’ reliance on the AI-CDS offset this impairment and increased performance to the level of their unaffected (control arm) counterparts^78^. Moreover, when given recommendations from the AI-CDS that differed from their initial decision, participants with lower confidence were more likely to switch their decision to that of the AI-CDS^42,61^.

## DISCUSSION

This study sought to clarify existing research on AI-CDS by identifying factors that influence the effectiveness of AI-assisted decision-making in medical settings and highlight promising factors for future study. However, of the three outcomes relevant to effectiveness that we looked at, clinician performance with AI (e.g., diagnostic accuracy) was much more commonly measured than clinician attitudes (e.g., trust) or clinician decisions (e.g., rates of agreement with AI-CDS recommendations). We note that measuring human-AI collaborative performance without assessing clinician-level decisions or attitudes such as trust leaves some uncertainty about reasons why collaborative performance improved or failed to do so. Despite this, only five studies measured all three of these outcomes. As a result, many of the factors identified have inconsistent impacts across studies, leaving much uncertainty about factors that may impact the use of AI-CDS.

Despite this uncertainty, there were four commonly studied factors that have clear impacts: clinician baseline attitude toward AI, clinician expertise, AI explainability, and AI correctness. Of these, baseline attitudes toward AI and AI correctness appeared to have the most consistent effects. Specifically, clinicians’ baseline attitudes toward AI (e.g., their trust in AI) guided their tendency to accept the AI-CDS’ recommendations (in 10/11 studies) and when clinicians were given incorrect advice from AI-CDS, their performance decreased (in 9/10 studies).

Consequently, we identified that when AI-CDS provides incorrect advice often enough and when clinicians’ trust in this AI-CDS is high, it has the potential to decrease clinicians’ performance relative to their baseline without AI assistance^57^. Because clinicians (of all experience levels) may not be able to identify incorrect AI advice, blindly increasing clinicians’ trust in AI recommendations can increase how often they defer to such advice and potentially decrease their decision-making performance.

This observation -that techniques to increase trust in AI may lead to over-reliance on AI-CDS-suggests the need for a paradigm shift; instead of aiming simply to increase trust in AI, a focus of much prior work^82–84^, researchers should aim for trust that is “just right,” promoting a level of trust that allows clinicians and health systems to benefit from AI-CDS, while avoiding the pitfalls of automation bias and over-reliance on AI^15,16,75^. This concept has been well-studied for non-AI technologies^33^, under the name of “appropriate trust”^85^ and has shown utility in medical and non-medical applications^14^. Under the concept of appropriate trust, clinicians learn the optimal level of trust they should place in AI-CDS relative to their own level of decision-making performance. In doing so, they aim to strike an ideal balance between under-utilization of AI and over-reliance on AI.

This framework of appropriate trust can be used to explain much of the heterogeneity in our current findings. Specifically, we found substantial heterogeneity in how clinicians’ level of expertise impacted AI-assisted decision-making performance (9/16 studies showing no relationship, 6/16 finding that non-experts benefit more, and 2/16 finding that experts benefit more). While expertise did not reliably predict which clinicians’ performance would increase from AI-CDS assistance, we found that more experienced clinicians (experts) were generally less trusting of AI-CDS and were less likely to accept AI recommendations^42,47,51,56^. Consequently, they were less likely to benefit from AI-CDS when it correctly provided advice that contradicted their own assessment. However, this also made experts more resistant to over-reliance (i.e., accepting incorrect AI advice) than their junior counterparts^39,51^. The synthesis of these observations is that expertise most directly predicts a clinicians’ tendency to accept AI recommendations. However, whether this difference in acceptance rates will increase clinicians’ decision-making performance is dependent on how well these clinicians perform relative to the AI-CDS; if the clinician performs well, then they likely only stand to decrease their performance by accepting AI advice. Similarly, if the clinician performs poorly on their own (as might be expected for non-experts), then higher trust in AI is beneficial as they have more room to improve from AI assistance. In the framework of appropriate trust, this is referred to as “trust calibration,”^33^ the concept that trust should be calibrated relative to the level of benefit that the AI-CDS stands to bring. In this respect, clinicians’ trust in AI can be considered well-calibrated with respect to clinical expertise as those who stand to benefit the least from AI-CDS also trust AI-CDS the least. In our review, this was highlighted most directly by Tschandl et al.,^42^ who identified that “if experts have high confidence in their initial diagnosis, they should ignore AI-based support or not use it at all”.

The concept of appropriate trust also enables us to analyze the heterogeneous results observed with respect to xAI. Specifically, our literature review shows that xAI often increases clinicians’ trust in AI-CDS but does not necessarily make this trust more appropriate. That is, xAI can increase clinicians’ acceptance of incorrect advice from the AI-CDS^68,69,72^ and when it does so often enough, this can harm human-AI collaborative performance^42,57^. This finding has been corroborated by recent non-medical research^86–89^. Moreover, a recent review of xAI identified that xAI can exacerbate certain cognitive biases, leading to decreases in decision-making accuracy^90^. However, we also identified conflicting evidence where xAI led to more appropriate trust. Specifically, xAI increased clinician acceptance of high-performing AI-CDS^64,68^ and decreased their trust in the AI-CDS when it provided incorrect recommendations^60,73^. Recent systematic reviews point out that these inconsistent findings may result from the lack of standard evaluation procedures for xAI^91,92^. In this context, our results highlight the potential for xAI to increase the appropriateness of clinician trust in AI and support the role of appropriate trust as a component in xAI evaluation frameworks.

This review additionally identified that studies focused on a narrow band of decision-making tasks in well-controlled environments. Specifically, the majority of studies assessed AI for diagnostic tasks (36/45), most AI systems were for some type of medical imaging (29/45). While AI has been very effective in medical imaging, results from these tasks may not generalize to others as image interpretation may be more dependent on search processes^93^ than clinical reasoning (e.g., for differential diagnosis) which may rely more on analytic and hypothesis-driven reasoning^94^. Additionally, only two out of forty-five studies conducted their study in the real-world, only one assessed changes in AI-assisted decision-making over time, many studies (12/45) used simulated AI systems that may not behave how real AI-CDS would and only three studies used large language models with interactive capabilities (the rest of provided probabilities or binary decisions). While these study design choices are reasonable, and maybe even preferred, when assessing how different factors impact AI-assisted decision-making, they have limited applicability as many of the assumptions made when conducting these studies will not hold in real-world environments. For example, it is well known that users of a technology will recalibrate their trust in that technology over time, often in response to feedback about the system’s performance^95^ or in response to changing environments^14^. However, only one study evaluated AI-assisted decision-making over time and no studies considered giving performance feedback to users. Several important questions emerge from these gaps such as: “are human-AI dynamics the same in real-world environments as they are in laboratory studies?”, “are there certain kinds of clinical tasks for which clinician trust in AI-CDS is well- or poorly-calibrated?”, and “what types of feedback about clinicians’ usage of AI-CDS enables users to calibrate their trust?”.

### Recommendations

To make sense of the heterogeneous results observed in existing AI-CDS research, we recommend that future research into AI-CDS should leverage the concept of appropriate trust^33,85^ to guide the design and evaluation of AI-CDS systems. From this framework, we offer the following four recommendations:

1. Researchers should measure: (1) clinicians’ attitudes towards the AI system (e.g., trust), (2) clinicians’ actions when presented with the AI-CDS (e.g., their acceptance or non-acceptance of AI-CDS recommendations), and (3) performance of the clinicians alone, the AI alone, and the human-AI collaboration, as all three are necessary to understand the complete picture around AI-assisted decision-making in medicine.
2. Researchers should evaluate clinicians’ interactions with AI-CDS longitudinally to understand how their attitudes, actions, and performance evolve over time.
3. Researchers should investigate the effect of providing feedback to clinicians about their performance or actions when using AI-CDS.
4. Researchers should conduct observational, and eventually interventional, studies in real-world clinical environments to validate whether clinician interactions with AI-CDS observed in controlled environments will translate to clinical practice.

### Limitations

Our review has several limitations. First, as with all literature reviews, some literature may have been missed during our search. Second, the current literature on AI-CDS focused largely on simple decisions and AI-CDS without much interactive capability. Because copilot-style AI systems^6,10^ bring unique interactive capabilities beyond the AI systems studied in current research, future work must be done to understand human-AI collaboration with these more dynamic AI systems. Third, our review required that users directly interact with an AI system in an experimental or in a real-world clinical setting, as this is the only way decision-making could be reliably studied. However, this omitted a substantial body of qualitative research about the design of AI interfaces, users’ experiences with the system, and sociotechnical issues potentially arising from the AI system that may impact clinicians’ attitudes towards AI. Fourth, we excluded a considerable number of studies with fewer than 30 participants as estimates derived from very small sample sizes are unlikely to be robust at the level of individual participants, and generalizability may also be limited by the limited range of participants. Nevertheless, this decision may have excluded some relevant research.

### Conclusion

This scoping review identified factors that influence AI-assisted decision-making. The most robust findings in our study were that clinicians’ attitudes towards AI (e.g., trust) consistently predicted their acceptance of AI recommendations and that when AI provided incorrect recommendations, clinicians’ decision-making performance decreased substantially. We integrated these findings with theoretical frameworks and used the concept of appropriate trust to provide recommendations for more robust research into AI-assisted medical decision-making.

We used this framework to analyze why the most commonly studied factors in our review - explainable AI and clinicians’ level of expertise - had heterogeneous impacts on decision-making performance. In this context, we found signals that clinicians’ trust in AI may be naturally appropriate – at a broad cohort level more experienced (typically higher-performing) clinicians were less trusting of AI-CDS than their junior counterparts, who stand to benefit more from AI assistance. However, trust may not be appropriate at the individual level as clinicians often could not reliably distinguish between correct recommendations which should be trusted and incorrect ones that should not. Similarly, explainable AI showed promise as it often increased clinicians’ trust in highly effective AI systems and reduced trust in error-prone ones. However, the benefits of xAI were not universally observed; xAI occasionally increased acceptance of incorrect advice. We conclude that the concept of appropriate trust is an essential tool for understanding the complex interactions between clinicians and AI-CDS systems and that future research should leverage this concept to facilitate more robust study of AI-assisted decision-making in medicine.

## Data Availability

The minimum necessary materials to generate results presented in this paper (i.e., included studies and relevant data attributes) are contained in the Supplementary Materials. The full set of studies, both included and excluded, as well as all extracted information, and the code used to generate figures and complete the analysis are available in an Open Science Framework repository (https://osf.io/un32b/).

## Code Availability

The code necessary to reproduce the analysis in this paper and generate figures, along with a readme file is available in an Open Science Framework repository (https://osf.io/un32b/).

## Author Contributions

NJJ, KEB, MRC, JSA contributed to the study design. NJJ conducted the literature search, and NJJ, KEB, RM, MM, and BC screened articles. NJJ and KEB processed the data. NJJ drafted the initial manuscript. NJJ, KEB, MRC, BC, LLN, NCB, JSA contributed to data analysis and interpretation, and reviewed the manuscript. NJJ and JSA were responsible for integrating revisions and finalizing the manuscript.

