## Supplementary Materials for "Factors Influencing the Effectiveness of AI-Assisted Decision-Making in Medicine: A Scoping Review" (from another version of this preprint: version 1, 2025-09-04)

### **Supplementary Table 1. PRISCMA-ScR Checklist**

**Preferred Reporting Items for Systematic reviews and Meta-Analyses extension for Scoping Reviews (PRISMA-ScR) Checklist**

| **SECTION** | **ITEM** | **PRISMA-ScR CHECKLIST ITEM** | **REPORTED ON PAGE #** |
| --- | --- | --- | --- |
| **TITLE** | | | |
| Title | 1 | Identify the report as a scoping review. | 1 |
| **ABSTRACT** | | | |
| Structured summary | 2 | Provide a structured summary that includes (as applicable): background, objectives, eligibility criteria, sources of evidence, charting methods, results, and conclusions that relate to the review questions and objectives. | 2 |
| **INTRODUCTION** | | | |
| Rationale | 3 | Describe the rationale for the review in the context of what is already known. Explain why the review questions/objectives lend themselves to a scoping review approach. | 4 |
| Objectives | 4 | Provide an explicit statement of the questions and objectives being addressed with reference to their key elements (e.g., population or participants, concepts, and context) or other relevant key elements used to conceptualize the review questions and/or objectives. | 5 |
| **METHODS** | | | |
| Protocol and registration | 5 | Indicate whether a review protocol exists; state if and where it can be accessed (e.g., a Web address); and if available, provide registration information, including the registration number. | 7 |
| Eligibility criteria | 6 | Specify characteristics of the sources of evidence used as eligibility criteria (e.g., years considered, language, and publication status), and provide a rationale. | 6 |
| Information sources* | 7 | Describe all information sources in the search (e.g., databases with dates of coverage and contact with authors to identify additional sources), as well as the date the most recent search was executed. | 5 |
| Search | 8 | Present the full electronic search strategy for at least 1 database, including any limits used, such that it could be repeated. | 5-6, Supplementary Materials p8 |
| Selection of sources of evidence† | 9 | State the process for selecting sources of evidence (i.e., screening and eligibility) included in the scoping review. | 6-7 |
| Data charting process‡ | 10 | Describe the methods of charting data from the included sources of evidence (e.g., calibrated forms or forms that have been tested by the team before their use, and whether data charting was done independently or in duplicate) and any processes for obtaining and confirming data from investigators. | 7 |
| Data items | 11 | List and define all variables for which data were sought and any assumptions and simplifications made. | 7 |
| Critical appraisal of individual sources of evidence§ | 12 | If done, provide a rationale for conducting a critical appraisal of included sources of evidence; describe the methods used and how this information was used in any data synthesis (if appropriate). | N/A |
| Synthesis of results | 13 | Describe the methods of handling and summarizing the data that were charted. | 7, OSF Repository (see README.txt) |
| **RESULTS** | | | |
| Selection of sources of evidence | 14 | Give numbers of sources of evidence screened, assessed for eligibility, and included in the review, with reasons for exclusions at each stage, ideally using a flow diagram. | 7 |
| Characteristics of sources of evidence | 15 | For each source of evidence, present characteristics for which data were charted and provide the citations. | Supplementary Materials p4. OSF Repository (Extracted_Processed.csv, Extracted_Factors_Results.csv) |
| Critical appraisal within sources of evidence | 16 | If done, present data on critical appraisal of included sources of evidence (see item 12). | N/A |
| Results of individual sources of evidence | 17 | For each included source of evidence, present the relevant data that were charted that relate to the review questions and objectives. | OSF Repository (Extracted_Processed.csv, Extracted_Factors_Results.csv) |
| Synthesis of results | 18 | Summarize and/or present the charting results as they relate to the review questions and objectives. | 7-13 |
| **DISCUSSION** | | | |
| Summary of evidence | 19 | Summarize the main results (including an overview of concepts, themes, and types of evidence available), link to the review questions and objectives, and consider the relevance to key groups. | 13-16 |
| Limitations | 20 | Discuss the limitations of the scoping review process. | 17 |
| Conclusions | 21 | Provide a general interpretation of the results with respect to the review questions and objectives, as well as potential implications and/or next steps. | 18 |
| **FUNDING** | | | |
| Funding | 22 | Describe sources of funding for the included sources of evidence, as well as sources of funding for the scoping review. Describe the role of the funders of the scoping review. | 20 |

JBI = Joanna Briggs Institute; PRISMA-ScR = Preferred Reporting Items for Systematic reviews and Meta-Analyses extension for Scoping Reviews.

* Where *sources of evidence* (see second footnote) are compiled from, such as bibliographic databases, social media platforms, and Web sites.

† A more inclusive/heterogeneous term used to account for the different types of evidence or data sources (e.g., quantitative and/or qualitative research, expert opinion, and policy documents) that may be eligible in a scoping review as opposed to only studies. This is not to be confused with *information sources* (see first footnote).

‡ The frameworks by Arksey and O’Malley (6) and Levac and colleagues (7) and the JBI guidance (4, 5) refer to the process of data extraction in a scoping review as data charting*.*

§ The process of systematically examining research evidence to assess its validity, results, and relevance before using it to inform a decision. This term is used for items 12 and 19 instead of "risk of bias" (which is more applicable to systematic reviews of interventions) to include and acknowledge the various sources of evidence that may be used in a scoping review (e.g., quantitative and/or qualitative research, expert opinion, and policy document).

*From:* Tricco AC, Lillie E, Zarin W, O'Brien KK, Colquhoun H, Levac D, et al. PRISMA Extension for Scoping Reviews (PRISMAScR): Checklist and Explanation. Ann Intern Med. 2018;169:467–473. [doi: 10.7326/M18-0850](http://annals.org/aim/fullarticle/2700389/prisma-extension-scoping-reviews-prisma-scr-checklist-explanation).

### **Queries used for study identification**

- This query was generated programmatically and used to search PubMed. The queries used to search Embase and WebOfScience are identical, but were changed to fit the syntax of their respective sites.
- For brevity, the queries used to search Embase and Web Of Science are located in the OSF Repository (<https://osf.io/un32b/?view_only=efbeaf42a9ef4aaab45a2cde69450265>) under “Lit_Review_Queries.txt”.

(("artificial intelligence"[tiab] OR "machine learning"[tiab] OR "deep learning"[tiab] OR "artificial-intelligence"[tiab] OR "machine-learning"[tiab] OR "deep-learning"[tiab] OR "AI"[tiab]) AND (("reader study"[tiab] OR "reader studies"[tiab] OR "vignette"[tiab] OR "vignettes"[tiab] OR "user study"[tiab] OR "usability study"[tiab]) OR ("human-AI"[tiab] OR "clinician-AI"[tiab] OR "physician-AI"[tiab] OR "physician-machine"[tiab] OR "AI based recommendation"[tiab] OR "AI based recommendations"[tiab] OR "AI based assistance"[tiab] OR "AI based support"[tiab] OR "AI based advice"[tiab] OR "AI based decision support"[tiab] OR "AI based diagnostic systems"[tiab] OR "AI based diagnosis"[tiab] OR "AI-based recommendation"[tiab] OR "AI-based recommendations"[tiab] OR "AI-based assistance"[tiab] OR "AI-based support"[tiab] OR "AI-based advice"[tiab] OR "AI-based decision support"[tiab] OR "AI-based diagnostic systems"[tiab] OR "AI-based diagnosis"[tiab] OR "AI recommendation"[tiab] OR "AI recommendations"[tiab] OR "AI assistance"[tiab] OR "AI support"[tiab] OR "AI advice"[tiab] OR "AI decision support"[tiab] OR "AI diagnostic systems"[tiab] OR "AI diagnosis"[tiab] OR "AI-recommendation"[tiab] OR "AI-recommendations"[tiab] OR "AI-assistance"[tiab] OR "AI-support"[tiab] OR "AI-advice"[tiab] OR "AI-decision support"[tiab] OR "AI-diagnostic systems"[tiab] OR "AI-diagnosis"[tiab] OR "AI-generated recommendation"[tiab] OR "AI-generated recommendations"[tiab] OR "AI-generated assistance"[tiab] OR "AI-generated support"[tiab] OR "AI-generated advice"[tiab] OR "AI-generated decision support"[tiab] OR "AI-generated diagnostic systems"[tiab] OR "AI-generated diagnosis"[tiab] OR "AI generated recommendation"[tiab] OR "AI generated recommendations"[tiab] OR "AI generated assistance"[tiab] OR "AI generated support"[tiab] OR "AI generated advice"[tiab] OR "AI generated decision support"[tiab] OR "AI generated diagnostic systems"[tiab] OR "AI generated diagnosis"[tiab] OR "AI-aided recommendation"[tiab] OR "AI-aided recommendations"[tiab] OR "AI-aided assistance"[tiab] OR "AI-aided support"[tiab] OR "AI-aided advice"[tiab] OR "AI-aided decision support"[tiab] OR "AI-aided diagnostic systems"[tiab] OR "AI-aided diagnosis"[tiab] OR "AI aided recommendation"[tiab] OR "AI aided recommendations"[tiab] OR "AI aided assistance"[tiab] OR "AI aided support"[tiab] OR "AI aided advice"[tiab] OR "AI aided decision support"[tiab] OR "AI aided diagnostic systems"[tiab] OR "AI aided diagnosis"[tiab] OR "machine learning based recommendation"[tiab] OR "machine learning based recommendations"[tiab] OR "machine learning based assistance"[tiab] OR "machine learning based support"[tiab] OR "machine learning based advice"[tiab] OR "machine learning based decision support"[tiab] OR "machine learning based diagnostic systems"[tiab] OR "machine learning based diagnosis"[tiab] OR "machine learning-based recommendation"[tiab] OR "machine learning-based recommendations"[tiab] OR "machine learning-based assistance"[tiab] OR "machine learning-based support"[tiab] OR "machine learning-based advice"[tiab] OR "machine learning-based decision support"[tiab] OR "machine learning-based diagnostic systems"[tiab] OR "machine learning-based diagnosis"[tiab] OR "machine learning recommendation"[tiab] OR "machine learning recommendations"[tiab] OR "machine learning assistance"[tiab] OR "machine learning support"[tiab] OR "machine learning advice"[tiab] OR "machine learning decision support"[tiab] OR "machine learning diagnostic systems"[tiab] OR "machine learning diagnosis"[tiab] OR "machine learning-recommendation"[tiab] OR "machine learning-recommendations"[tiab] OR "machine learning-assistance"[tiab] OR "machine learning-support"[tiab] OR "machine learning-advice"[tiab] OR "machine learning-decision support"[tiab] OR "machine learning-diagnostic systems"[tiab] OR "machine learning-diagnosis"[tiab] OR "machine learning-generated recommendation"[tiab] OR "machine learning-generated recommendations"[tiab] OR "machine learning-generated assistance"[tiab] OR "machine learning-generated support"[tiab] OR "machine learning-generated advice"[tiab] OR "machine learning-generated decision support"[tiab] OR "machine learning-generated diagnostic systems"[tiab] OR "machine learning-generated diagnosis"[tiab] OR "machine learning generated recommendation"[tiab] OR "machine learning generated recommendations"[tiab] OR "machine learning generated assistance"[tiab] OR "machine learning generated support"[tiab] OR "machine learning generated advice"[tiab] OR "machine learning generated decision support"[tiab] OR "machine learning generated diagnostic systems"[tiab] OR "machine learning generated diagnosis"[tiab] OR "machine learning-aided recommendation"[tiab] OR "machine learning-aided recommendations"[tiab] OR "machine learning-aided assistance"[tiab] OR "machine learning-aided support"[tiab] OR "machine learning-aided advice"[tiab] OR "machine learning-aided decision support"[tiab] OR "machine learning-aided diagnostic systems"[tiab] OR "machine learning-aided diagnosis"[tiab] OR "machine learning aided recommendation"[tiab] OR "machine learning aided recommendations"[tiab] OR "machine learning aided assistance"[tiab] OR "machine learning aided support"[tiab] OR "machine learning aided advice"[tiab] OR "machine learning aided decision support"[tiab] OR "machine learning aided diagnostic systems"[tiab] OR "machine learning aided diagnosis"[tiab] OR "deep learning based recommendation"[tiab] OR "deep learning based recommendations"[tiab] OR "deep learning based assistance"[tiab] OR "deep learning based support"[tiab] OR "deep learning based advice"[tiab] OR "deep learning based decision support"[tiab] OR "deep learning based diagnostic systems"[tiab] OR "deep learning based diagnosis"[tiab] OR "deep learning-based recommendation"[tiab] OR "deep learning-based recommendations"[tiab] OR "deep learning-based assistance"[tiab] OR "deep learning-based support"[tiab] OR "deep learning-based advice"[tiab] OR "deep learning-based decision support"[tiab] OR "deep learning-based diagnostic systems"[tiab] OR "deep learning-based diagnosis"[tiab] OR "deep learning recommendation"[tiab] OR "deep learning recommendations"[tiab] OR "deep learning assistance"[tiab] OR "deep learning support"[tiab] OR "deep learning advice"[tiab] OR "deep learning decision support"[tiab] OR "deep learning diagnostic systems"[tiab] OR "deep learning diagnosis"[tiab] OR "deep learning-recommendation"[tiab] OR "deep learning-recommendations"[tiab] OR "deep learning-assistance"[tiab] OR "deep learning-support"[tiab] OR "deep learning-advice"[tiab] OR "deep learning-decision support"[tiab] OR "deep learning-diagnostic systems"[tiab] OR "deep learning-diagnosis"[tiab] OR "deep learning-generated recommendation"[tiab] OR "deep learning-generated recommendations"[tiab] OR "deep learning-generated assistance"[tiab] OR "deep learning-generated support"[tiab] OR "deep learning-generated advice"[tiab] OR "deep learning-generated decision support"[tiab] OR "deep learning-generated diagnostic systems"[tiab] OR "deep learning-generated diagnosis"[tiab] OR "deep learning generated recommendation"[tiab] OR "deep learning generated recommendations"[tiab] OR "deep learning generated assistance"[tiab] OR "deep learning generated support"[tiab] OR "deep learning generated advice"[tiab] OR "deep learning generated decision support"[tiab] OR "deep learning generated diagnostic systems"[tiab] OR "deep learning generated diagnosis"[tiab] OR "deep learning-aided recommendation"[tiab] OR "deep learning-aided recommendations"[tiab] OR "deep learning-aided assistance"[tiab] OR "deep learning-aided support"[tiab] OR "deep learning-aided advice"[tiab] OR "deep learning-aided decision support"[tiab] OR "deep learning-aided diagnostic systems"[tiab] OR "deep learning-aided diagnosis"[tiab] OR "deep learning aided recommendation"[tiab] OR "deep learning aided recommendations"[tiab] OR "deep learning aided assistance"[tiab] OR "deep learning aided support"[tiab] OR "deep learning aided advice"[tiab] OR "deep learning aided decision support"[tiab] OR "deep learning aided diagnostic systems"[tiab] OR "deep learning aided diagnosis"[tiab] OR "artificial intelligence based recommendation"[tiab] OR "artificial intelligence based recommendations"[tiab] OR "artificial intelligence based assistance"[tiab] OR "artificial intelligence based support"[tiab] OR "artificial intelligence based advice"[tiab] OR "artificial intelligence based decision support"[tiab] OR "artificial intelligence based diagnostic systems"[tiab] OR "artificial intelligence based diagnosis"[tiab] OR "artificial intelligence-based recommendation"[tiab] OR "artificial intelligence-based recommendations"[tiab] OR "artificial intelligence-based assistance"[tiab] OR "artificial intelligence-based support"[tiab] OR "artificial intelligence-based advice"[tiab] OR "artificial intelligence-based decision support"[tiab] OR "artificial intelligence-based diagnostic systems"[tiab] OR "artificial intelligence-based diagnosis"[tiab] OR "artificial intelligence recommendation"[tiab] OR "artificial intelligence recommendations"[tiab] OR "artificial intelligence assistance"[tiab] OR "artificial intelligence support"[tiab] OR "artificial intelligence advice"[tiab] OR "artificial intelligence decision support"[tiab] OR "artificial intelligence diagnostic systems"[tiab] OR "artificial intelligence diagnosis"[tiab] OR "artificial intelligence-recommendation"[tiab] OR "artificial intelligence-recommendations"[tiab] OR "artificial intelligence-assistance"[tiab] OR "artificial intelligence-support"[tiab] OR "artificial intelligence-advice"[tiab] OR "artificial intelligence-decision support"[tiab] OR "artificial intelligence-diagnostic systems"[tiab] OR "artificial intelligence-diagnosis"[tiab] OR "artificial intelligence-generated recommendation"[tiab] OR "artificial intelligence-generated recommendations"[tiab] OR "artificial intelligence-generated assistance"[tiab] OR "artificial intelligence-generated support"[tiab] OR "artificial intelligence-generated advice"[tiab] OR "artificial intelligence-generated decision support"[tiab] OR "artificial intelligence-generated diagnostic systems"[tiab] OR "artificial intelligence-generated diagnosis"[tiab] OR "artificial intelligence generated recommendation"[tiab] OR "artificial intelligence generated recommendations"[tiab] OR "artificial intelligence generated assistance"[tiab] OR "artificial intelligence generated support"[tiab] OR "artificial intelligence generated advice"[tiab] OR "artificial intelligence generated decision support"[tiab] OR "artificial intelligence generated diagnostic systems"[tiab] OR "artificial intelligence generated diagnosis"[tiab] OR "artificial intelligence-aided recommendation"[tiab] OR "artificial intelligence-aided recommendations"[tiab] OR "artificial intelligence-aided assistance"[tiab] OR "artificial intelligence-aided support"[tiab] OR "artificial intelligence-aided advice"[tiab] OR "artificial intelligence-aided decision support"[tiab] OR "artificial intelligence-aided diagnostic systems"[tiab] OR "artificial intelligence-aided diagnosis"[tiab] OR "artificial intelligence aided recommendation"[tiab] OR "artificial intelligence aided recommendations"[tiab] OR "artificial intelligence aided assistance"[tiab] OR "artificial intelligence aided support"[tiab] OR "artificial intelligence aided advice"[tiab] OR "artificial intelligence aided decision support"[tiab] OR "artificial intelligence aided diagnostic systems"[tiab] OR "artificial intelligence aided diagnosis"[tiab] OR "ML based recommendation"[tiab] OR "ML based recommendations"[tiab] OR "ML based assistance"[tiab] OR "ML based support"[tiab] OR "ML based advice"[tiab] OR "ML based decision support"[tiab] OR "ML based diagnostic systems"[tiab] OR "ML based diagnosis"[tiab] OR "ML-based recommendation"[tiab] OR "ML-based recommendations"[tiab] OR "ML-based assistance"[tiab] OR "ML-based support"[tiab] OR "ML-based advice"[tiab] OR "ML-based decision support"[tiab] OR "ML-based diagnostic systems"[tiab] OR "ML-based diagnosis"[tiab] OR "ML recommendation"[tiab] OR "ML recommendations"[tiab] OR "ML assistance"[tiab] OR "ML support"[tiab] OR "ML advice"[tiab] OR "ML decision support"[tiab] OR "ML diagnostic systems"[tiab] OR "ML diagnosis"[tiab] OR "ML-recommendation"[tiab] OR "ML-recommendations"[tiab] OR "ML-assistance"[tiab] OR "ML-support"[tiab] OR "ML-advice"[tiab] OR "ML-decision support"[tiab] OR "ML-diagnostic systems"[tiab] OR "ML-diagnosis"[tiab] OR "ML-generated recommendation"[tiab] OR "ML-generated recommendations"[tiab] OR "ML-generated assistance"[tiab] OR "ML-generated support"[tiab] OR "ML-generated advice"[tiab] OR "ML-generated decision support"[tiab] OR "ML-generated diagnostic systems"[tiab] OR "ML-generated diagnosis"[tiab] OR "ML generated recommendation"[tiab] OR "ML generated recommendations"[tiab] OR "ML generated assistance"[tiab] OR "ML generated support"[tiab] OR "ML generated advice"[tiab] OR "ML generated decision support"[tiab] OR "ML generated diagnostic systems"[tiab] OR "ML generated diagnosis"[tiab] OR "ML-aided recommendation"[tiab] OR "ML-aided recommendations"[tiab] OR "ML-aided assistance"[tiab] OR "ML-aided support"[tiab] OR "ML-aided advice"[tiab] OR "ML-aided decision support"[tiab] OR "ML-aided diagnostic systems"[tiab] OR "ML-aided diagnosis"[tiab] OR "ML aided recommendation"[tiab] OR "ML aided recommendations"[tiab] OR "ML aided assistance"[tiab] OR "ML aided support"[tiab] OR "ML aided advice"[tiab] OR "ML aided decision support"[tiab] OR "ML aided diagnostic systems"[tiab] OR "ML aided diagnosis"[tiab] OR "DL based recommendation"[tiab] OR "DL based recommendations"[tiab] OR "DL based assistance"[tiab] OR "DL based support"[tiab] OR "DL based advice"[tiab] OR "DL based decision support"[tiab] OR "DL based diagnostic systems"[tiab] OR "DL based diagnosis"[tiab] OR "DL-based recommendation"[tiab] OR "DL-based recommendations"[tiab] OR "DL-based assistance"[tiab] OR "DL-based support"[tiab] OR "DL-based advice"[tiab] OR "DL-based decision support"[tiab] OR "DL-based diagnostic systems"[tiab] OR "DL-based diagnosis"[tiab] OR "DL recommendation"[tiab] OR "DL recommendations"[tiab] OR "DL assistance"[tiab] OR "DL support"[tiab] OR "DL advice"[tiab] OR "DL decision support"[tiab] OR "DL diagnostic systems"[tiab] OR "DL diagnosis"[tiab] OR "DL-recommendation"[tiab] OR "DL-recommendations"[tiab] OR "DL-assistance"[tiab] OR "DL-support"[tiab] OR "DL-advice"[tiab] OR "DL-decision support"[tiab] OR "DL-diagnostic systems"[tiab] OR "DL-diagnosis"[tiab] OR "DL-generated recommendation"[tiab] OR "DL-generated recommendations"[tiab] OR "DL-generated assistance"[tiab] OR "DL-generated support"[tiab] OR "DL-generated advice"[tiab] OR "DL-generated decision support"[tiab] OR "DL-generated diagnostic systems"[tiab] OR "DL-generated diagnosis"[tiab] OR "DL generated recommendation"[tiab] OR "DL generated recommendations"[tiab] OR "DL generated assistance"[tiab] OR "DL generated support"[tiab] OR "DL generated advice"[tiab] OR "DL generated decision support"[tiab] OR "DL generated diagnostic systems"[tiab] OR "DL generated diagnosis"[tiab] OR "DL-aided recommendation"[tiab] OR "DL-aided recommendations"[tiab] OR "DL-aided assistance"[tiab] OR "DL-aided support"[tiab] OR "DL-aided advice"[tiab] OR "DL-aided decision support"[tiab] OR "DL-aided diagnostic systems"[tiab] OR "DL-aided diagnosis"[tiab] OR "DL aided recommendation"[tiab] OR "DL aided recommendations"[tiab] OR "DL aided assistance"[tiab] OR "DL aided support"[tiab] OR "DL aided advice"[tiab] OR "DL aided decision support"[tiab] OR "DL aided diagnostic systems"[tiab] OR "DL aided diagnosis"[tiab] OR "model based recommendation"[tiab] OR "model based recommendations"[tiab] OR "model based assistance"[tiab] OR "model based support"[tiab] OR "model based advice"[tiab] OR "model based decision support"[tiab] OR "model based diagnostic systems"[tiab] OR "model based diagnosis"[tiab] OR "model-based recommendation"[tiab] OR "model-based recommendations"[tiab] OR "model-based assistance"[tiab] OR "model-based support"[tiab] OR "model-based advice"[tiab] OR "model-based decision support"[tiab] OR "model-based diagnostic systems"[tiab] OR "model-based diagnosis"[tiab] OR "model recommendation"[tiab] OR "model recommendations"[tiab] OR "model assistance"[tiab] OR "model support"[tiab] OR "model advice"[tiab] OR "model decision support"[tiab] OR "model diagnostic systems"[tiab] OR "model diagnosis"[tiab] OR "model-recommendation"[tiab] OR "model-recommendations"[tiab] OR "model-assistance"[tiab] OR "model-support"[tiab] OR "model-advice"[tiab] OR "model-decision support"[tiab] OR "model-diagnostic systems"[tiab] OR "model-diagnosis"[tiab] OR "model-generated recommendation"[tiab] OR "model-generated recommendations"[tiab] OR "model-generated assistance"[tiab] OR "model-generated support"[tiab] OR "model-generated advice"[tiab] OR "model-generated decision support"[tiab] OR "model-generated diagnostic systems"[tiab] OR "model-generated diagnosis"[tiab] OR "model generated recommendation"[tiab] OR "model generated recommendations"[tiab] OR "model generated assistance"[tiab] OR "model generated support"[tiab] OR "model generated advice"[tiab] OR "model generated decision support"[tiab] OR "model generated diagnostic systems"[tiab] OR "model generated diagnosis"[tiab] OR "model-aided recommendation"[tiab] OR "model-aided recommendations"[tiab] OR "model-aided assistance"[tiab] OR "model-aided support"[tiab] OR "model-aided advice"[tiab] OR "model-aided decision support"[tiab] OR "model-aided diagnostic systems"[tiab] OR "model-aided diagnosis"[tiab] OR "model aided recommendation"[tiab] OR "model aided recommendations"[tiab] OR "model aided assistance"[tiab] OR "model aided support"[tiab] OR "model aided advice"[tiab] OR "model aided decision support"[tiab] OR "model aided diagnostic systems"[tiab] OR "model aided diagnosis"[tiab] OR "computer based recommendation"[tiab] OR "computer based recommendations"[tiab] OR "computer based assistance"[tiab] OR "computer based support"[tiab] OR "computer based advice"[tiab] OR "computer based decision support"[tiab] OR "computer based diagnostic systems"[tiab] OR "computer based diagnosis"[tiab] OR "computer-based recommendation"[tiab] OR "computer-based recommendations"[tiab] OR "computer-based assistance"[tiab] OR "computer-based support"[tiab] OR "computer-based advice"[tiab] OR "computer-based decision support"[tiab] OR "computer-based diagnostic systems"[tiab] OR "computer-based diagnosis"[tiab] OR "computer recommendation"[tiab] OR "computer recommendations"[tiab] OR "computer assistance"[tiab] OR "computer support"[tiab] OR "computer advice"[tiab] OR "computer decision support"[tiab] OR "computer diagnostic systems"[tiab] OR "computer diagnosis"[tiab] OR "computer-recommendation"[tiab] OR "computer-recommendations"[tiab] OR "computer-assistance"[tiab] OR "computer-support"[tiab] OR "computer-advice"[tiab] OR "computer-decision support"[tiab] OR "computer-diagnostic systems"[tiab] OR "computer-diagnosis"[tiab] OR "computer-generated recommendation"[tiab] OR "computer-generated recommendations"[tiab] OR "computer-generated assistance"[tiab] OR "computer-generated support"[tiab] OR "computer-generated advice"[tiab] OR "computer-generated decision support"[tiab] OR "computer-generated diagnostic systems"[tiab] OR "computer-generated diagnosis"[tiab] OR "computer generated recommendation"[tiab] OR "computer generated recommendations"[tiab] OR "computer generated assistance"[tiab] OR "computer generated support"[tiab] OR "computer generated advice"[tiab] OR "computer generated decision support"[tiab] OR "computer generated diagnostic systems"[tiab] OR "computer generated diagnosis"[tiab] OR "computer-aided recommendation"[tiab] OR "computer-aided recommendations"[tiab] OR "computer-aided assistance"[tiab] OR "computer-aided support"[tiab] OR "computer-aided advice"[tiab] OR "computer-aided decision support"[tiab] OR "computer-aided diagnostic systems"[tiab] OR "computer aided recommendation"[tiab] OR "computer aided recommendations"[tiab] OR "computer aided assistance"[tiab] OR "computer aided support"[tiab] OR "computer aided advice"[tiab] OR "computer aided decision support"[tiab] OR "computer aided diagnostic systems"[tiab] OR "diagnostic assistance"[tiab] OR "diagnostic advice"[tiab] OR "decision-aid"[tiab] OR "decision-aids"[tiab] OR "diagnostic aid"[tiab] OR "diagnostic aids"[tiab] OR "automated diagnosis"[tiab] OR "automated diagnoses"[tiab] OR "decision support system"[tiab])) AND ("medicine"[tiab] OR "doctor"[tiab] OR "physician"[tiab] OR "clinician"[tiab] OR "clinic"[tiab] OR "clinics"[tiab] OR "hospital"[tiab] OR "hospitals"[tiab] OR "diagnose"[tiab] OR "diagnosis"[tiab] OR "diagnoses"[tiab] OR "health"[tiab] OR "healthcare"[tiab] OR "patient"[tiab] OR "patients"[tiab])) NOT "review"[title]
